# High Density Lipoprotein pathway as a therapeutic target for coronary heart disease: individual participant meta-analysis in 28,597 individuals with 4197 coronary events

**DOI:** 10.1101/2020.03.02.19010173

**Authors:** Amy R Mulick, David Prieto-Merino, Therese Tillin, Aki Havulinna, Martin Shipley, Desiree Valera-Gran, Aleksandra Gentry-Maharaj, Andy Ryan, Meena Kumari, J Wouter Jukema, Alex McConnachie, Veikko Salomaa, Nish Chaturvedi, Goya Wannamethee, Usha Menon, Barbara Jefferis, Mika Kivimaki, Chris J Packard, Naveed Sattar, John Whittaker, Aroon Hingorani, George B Ploubidis, JP Casas

## Abstract

**Importance:** Cholesterol content in high-density lipoprotein particles (HDL-C) is associated inversely with coronary heart disease (CHD), but findings from Mendelian randomization studies and randomized trials of HDL-C raising drugs have questioned whether this link is causal. However, these analyses do not exclude a causal role for specific HDL sub-fractions of different density, mobility, size and composition.

**Objective:** To determine whether sub-components of the HDL pathway exhibit differing relationships with CHD risk.

*Design:* In seven longitudinal studies, we used factor analysis to reduce 21 measures of HDL particle size and lipid content to a smaller number of factors representing different components of the HDL pathway. We constructed factor scores and modelled their associations on CHD risk in adjusted Cox regression analyses. We pooled results using random-effects meta-analysis.

**Setting:** Seven population-, individual-, occupational- or community-based longitudinal studies in the UK and Finland.

**Participants:** 28,597 participants (49% female, mean age 59.6 years) contributed to the analysis.

**Exposures:** Sub-components of the HDL pathway, characterized by 21 measures of HDL size and lipid content based on nuclear magnetic resonance spectroscopy.

**Main Outcomes:** Incident fatal or non-fatal CHD.

**Results:** We identified 4 HDL components with highly replicable across studies; 3 were indices of particle size/composition (extra-large (XL), large (L) and medium/small (MS)), and the other an index of triglycerides (TG) carried in HDL of all sizes. After up to 17 years of follow-up, 4179 incident CHD cases occurred. After adjusting for age, sex, ethnicity, smoking, systolic blood pressure, body mass index, diabetes and LDL-C, higher levels of the XL and MS factors were linked to a reduced risk of CHD (hazard ratio per 1 standard deviation (SD) increase 0.88 [95% CI 0.85, 0.92] and 0.91 [0.87, 0.94]). In contrast, a SD increase in the level of the TG factor was associated with increased risk of CHD (1.10 [1.07, 1.14]).

**Conclusions and Relevance:** We found qualitative differences between sub-components of the HDL pathway and the risk of developing CHD. Discovery of the biological determinants of these components, possibly through genetic analysis, will facilitate selection of drug targets and inform trial design.

**Key Points:** *Question:* Can investigation of sub-components of the high-density lipoprotein (HDL) pathway, measured through nuclear magnetic resonance spectroscopy, point to specific therapeutic targets for prevention of coronary heart disease (CHD)?

*Findings:* Using individual-level data from seven longitudinal studies including 28,597 participants and 4197 CHD events, we identified two components of the HDL pathway that were associated with reduced, and one that was associated with increased, risk of CHD.

*Meaning:* These sub-components of the HDL pathway, if causally related to atherogenesis, offer a route to more precise therapeutic targets for prevention of CHD.

## Introduction

There is extensive epidemiological evidence that high levels of cholesterol in high density lipoproteins (HDL-C) is associated with reduced risk of cardiovascular events^1^. These observations led to a drug-discovery pipeline which employed HDL-C as target, and this biomarker informed compound selection and trial design^2^. As a result, several drugs that raise HDL-C by the modulation of several protein-targets were identified that were expected to decrease the risk of cardiovascular events. However, randomized trials of drugs to raise HDL-C yielded inconsistent results^3-6^; for example the CETP inhibitor anacetrapib^4^ was shown to reduce coronary heart disease (CHD) risk, while niacin^5^, which targets a different protein, had no discernible effect. This suggests that HDL-C is incapable of distinguishing which protein is a valid therapeutic target for CHD prevention.

Some^7^, but not all^8^, Mendelian randomization studies designed to ascertain causality of HDL-C (instead of the protein target of HDL-C raising drugs), suggest that HDL-C is unlikely to be causally related to coronary atherosclerosis. However, evidence from genome-wide association studies indicates that variants at certain HDL-C raising loci do exhibit associations with CHD^9, 10^, whilst others do not, again suggesting that HDL-C is incapable of distinguishing which gene-encoded proteins are valid therapeutic targets for CHD prevention.

Measurement of cholesterol content in HDL particles alone may not capture the full complexity of the HDL pathway, which is composed of a heterogeneous family of lipoprotein particles of different density, mobility, size and composition that are under the control of different enzymatic processes^11^, which may have diverse biological functions and hence a differential role in the origins of cardiovascular events^12^. In support of this is the finding that rare genetic disorders of enzymes that control HDL metabolism seem to affect different sub-classes of HDL particles and this is associated with different phenotypes^13^. Moreover, some recent assays that measure HDL-C efflux capacity have shown this to be associated with CHD, independently of HDL-C^14, 15^.

To quantify the associations of specific components of the HDL pathway in the development of CHD, we used Nuclear Magnetic Resonance (NMR) spectroscopy^16^ to measure 21 biomarkers encompassing lipoprotein size and composition in seven longitudinal studies with 28,597 participants and 4197 coronary events.

## Methods

### Studies contributing data to this analysis

Data from the British Regional Heart Study (BRHS), British Women’s Heart and Health Study (BWHHS), FINRISK, PROspective Study of Pravastatin in the Elderly at Risk (PROSPER), Southall And Brent REvisited (SABRE), United Kingdom Collaborative Trial of Ovarian Cancer Screening (UKCTOCS) and the Whitehall-II study (WHII) were used for this investigation. Study details have previously been described^17-23^ and are also summarized in **Online-only material**. Figure 1 presents the flow of participants selected for these analyses. Study design, outcome definition, and time of blood sampling used for NMR spectroscopy is described in **eTable 1**.

**Table 1:** Baseline subject demographics for CHD regression analyses (n=28,597). All numbers are mean (SD) or n (%).

|  | **BRHS**<br>**(n=3535)** | **BWHHS**<br>**(n=3391)** | **FINRISK**<br>**(n=7328)** | **PROSPER**<br>**(n=3071)** | **SABRE**<br>**(n=2644)** | **UKCTOCS**<br>**(n=3163)** | **WHII**<br>**(n=5465)** |
| --- | --- | --- | --- | --- | --- | --- | --- |
| **Age (years)** | 68.6 (5.49) | 68.64 (5.48) | 47.77 (13) | 75.6 (3.32) | 51.92 (7.08) | 64.73 (6.2) | 55.56 (6.01) |
| **Gender (female)** | 0 (0%) | 3391 (100%) | 3754 (51%) | 1770 (58%) | 388 (15%) | 3163 (100%) | 1561 (29%) |
| **Ethnicity (white)** | 3512 (99%) | 3378 (>99%) | 7328 (100%) | 3071 (100%) | 1246 (47%) | 3068 (97%) | 5101 (93%) |
| **Smoking (current)** | 455 (13%) | 369 (11%) | 1740 (24%) | 934 (30%) | 588 (22%) | NA | 541 (9.9%) |
| **Systolic BP (mmHg)** | 149.7 (23.7) | 153.2 (25.3) | 135.7 (19.9) | 155.9 (22) | 124.3 (17.5) | NA | 123 (16.4) |
| **BMI (kg/m<sup>2</sup>)** | 26.84 (3.63) | 27.4 (4.82) | 26.61 (4.52) | 29.28 (146) | 26.12 (3.77) | 27.02 (5.37) | 26.03 (3.88) |
| **<25** | 1100 (31%) | 1295 (38%) | 2897 (40%) | 1139 (37%) | 1082 (41%) | 1273 (40%) | 2049 (37%) |
| **25-30** | 1869 (53%) | 1338 (39%) | 3012 (41%) | 1323 (43%) | 1220 (46%) | 1164 (37%) | 2061 (38%) |
| **>30** | 565 (16%) | 757 (22%) | 1418 (19%) | 608 (20%) | 341 (13%) | 740 (23%) | 640 (12%) |
| **Diabetes(yes)** | 205 (5.8%) | 302 (8.9%) | 393 (5.4%) | 245 (8%) | 303 (11%) | NA | 264 (4.8%) |
| **Total cholesterol (mmol/l)\*** | 6.026 (1.07) | 6.655 (1.18) | 5.547 (1.06) | 5.033 (1.08) | 6.004 (1.1) | NA | 5.942 (1.04) |
| **NMR Total cholesterol (mmol/l)** | 4.532 (0.92) | 6.011 (1.33) | 5.329 (1.11) | 4.218 (0.951) | 3.958 (0.905) | 4.94 (1.05) | 5.071 (0.909) |
| **HDL cholesterol (mmol/l)\*** | 1.329 (0.341) | 1.665 (0.455) | 1.401 (0.362) | 1.397 (0.388) | 1.29 (0.365) | NA | 1.464 (0.393) |
| **NMR HDL cholesterol (mmol/l)** | 1.197 (0.306) | 1.676 (0.451) | 1.581 (0.389) | 1.359 (0.279) | 1.041 (0.234) | 1.624 (0.411) | 1.514 (0.329) |
| **LDL cholesterol (mmol/l)\*** | 3.893 (0.963) | 4.165 (1.07) | 3.485 (0.933) | 3.125 (0.974) | 3.92 (0.964) | NA | 3.863 (0.933) |
| **NMR LDL cholesterol (mmol/l)** | 1.815 (0.515) | 2.436 (0.75) | 1.919 (0.602) | 1.388 (0.494) | 1.42 (0.453) | 1.803 (0.584) | 1.895 (0.486) |
| **Triglycerides (mmol/l)\*** | 1.827 (1.07) | 1.833 (0.952) | 1.483 (1.03) | 1.38 (0.654) | 1.888 (1.39) | NA | 1.355 (0.864) |
| **NMR Triglycerides (mmol/l)** | 1.561 (0.744) | 1.664 (0.84) | 1.306 (0.683) | 1.288 (0.56) | 1.006 (0.401) | 1.721 (0.895) | 1.241 (0.468) |
| **Non-HDL triglycerides (mmol/l)\*** | 1.404 (0.709) | 1.49 (0.798) | 1.132 (0.63) | 1.149 (0.527) | 0.8975 (0.377) | 1.529 (0.842) | 1.102 (0.441) |
| **On lipid-lowering medication\*\*** | 192 (5.4%) | 201 (5.9%) | 185 (2.5%) | 1503 (49%) | 4 (<1%) | NA | 119 (2.2%) |
| **Prior history of CHD** | 0 (0%) | 0 (0%) | 0 (0%) | 0 (0%) | 0 (0%) | NA | 0 (0%) |
| **Had CHD event** | 331 (9.4%) | 248 (7.3%) | 802 (11%) | 217 (7.1%) | 821 (31%) | 1565 (49%) | 195 (3.6%) |
| **Follow-up time (years)** | 8.57 (2.64) | 10.49 (2.7) | 13.87 (2.79) | 2.71 (0.58) | 17.26 (5.48) | 5.20 (2.27) | 6.23 (1.24) |

\*fasting measures; \*\*number in pravastatin arm of the PROSPER trial; NA, not available.

|  | BRHS (n=3535) | BWHHS (n=3391) | FINRISK (n=7328) | PROSPER (n=3071) | SABRE (n=2644) | UKCTOCS (n=3163) | WHII (n=5465) |
| --- | --- | --- | --- | --- | --- | --- | --- |
| **Age (years)** | 68.6 (5.49) | 68.64 (5.48) | 47.77 (13) | 75.6 (3.32) | 51.92 (7.08) | 64.73 (6.2) | 55.56 (6.01) |
| **Gender (female)** | 0 (0%) | 3391 (100%) | 3754 (51%) | 1770 (58%) | 388 (15%) | 3163 (100%) | 1561 (29%) |
| **Ethnicity (white)** | 3512 (99%) | 3378 (>99%) | 7328 (100%) | 3071 (100%) | 1246 (47%) | 3068 (97%) | 5101 (93%) |
| **Smoking (current)** | 455 (13%) | 369 (11%) | 1740 (24%) | 934 (30%) | 588 (22%) | NA | 541 (9.9%) |
| **Systolic BP (mmHg)** | 149.7 (23.7) | 153.2 (25.3) | 135.7 (19.9) | 155.9 (22) | 124.3 (17.5) | NA | 123 (16.4) |
| **BMI (kg/m<sup>2</sup>)** | 26.84 (3.63) | 27.4 (4.82) | 26.61 (4.52) | 29.28 (146) | 26.12 (3.77) | 27.02 (5.37) | 26.03 (3.88) |
| **<25** | 1100 (31%) | 1295 (38%) | 2897 (40%) | 1139 (37%) | 1082 (41%) | 1273 (40%) | 2049 (37%) |
| **25-30** | 1869 (53%) | 1338 (39%) | 3012 (41%) | 1323 (43%) | 1220 (46%) | 1164 (37%) | 2061 (38%) |
| **>30** | 565 (16%) | 757 (22%) | 1418 (19%) | 608 (20%) | 341 (13%) | 740 (23%) | 640 (12%) |
| **Diabetes(yes)** | 205 (5.8%) | 302 (8.9%) | 393 (5.4%) | 245 (8%) | 303 (11%) | NA | 264 (4.8%) |
| **Total cholesterol (mmol/l)\*** | 6.026 (1.07) | 6.655 (1.18) | 5.547 (1.06) | 5.033 (1.08) | 6.004 (1.1) | NA | 5.942 (1.04) |
| **NMR Total cholesterol (mmol/l)** | 4.532 (0.92) | 6.011 (1.33) | 5.329 (1.11) | 4.218 (0.951) | 3.958 (0.905) | 4.94 (1.05) | 5.071 (0.909) |
| **HDL cholesterol (mmol/l)\*** | 1.329 (0.341) | 1.665 (0.455) | 1.401 (0.362) | 1.397 (0.388) | 1.29 (0.365) | NA | 1.464 (0.393) |
| **NMR HDL cholesterol (mmol/l)** | 1.197 (0.306) | 1.676 (0.451) | 1.581 (0.389) | 1.359 (0.279) | 1.041 (0.234) | 1.624 (0.411) | 1.514 (0.329) |
| **LDL cholesterol (mmol/l)\*** | 3.893 (0.963) | 4.165 (1.07) | 3.485 (0.933) | 3.125 (0.974) | 3.92 (0.964) | NA | 3.863 (0.933) |
| **NMR LDL cholesterol (mmol/l)** | 1.815 (0.515) | 2.436 (0.75) | 1.919 (0.602) | 1.388 (0.494) | 1.42 (0.453) | 1.803 (0.584) | 1.895 (0.486) |
| **Triglycerides (mmol/l)\*** | 1.827 (1.07) | 1.833 (0.952) | 1.483 (1.03) | 1.38 (0.654) | 1.888 (1.39) | NA | 1.355 (0.864) |
| **NMR Triglycerides (mmol/l)** | 1.561 (0.744) | 1.664 (0.84) | 1.306 (0.683) | 1.288 (0.56) | 1.006 (0.401) | 1.721 (0.895) | 1.241 (0.468) |
| **Non-HDL triglycerides (mmol/l)\*** | 1.404 (0.709) | 1.49 (0.798) | 1.132 (0.63) | 1.149 (0.527) | 0.8975 (0.377) | 1.529 (0.842) | 1.102 (0.441) |
| **On lipid-lowering medication\*\*** | 192 (5.4%) | 201 (5.9%) | 185 (2.5%) | 1503 (49%) | 4 (<1%) | NA | 119 (2.2%) |
| **Prior history of CHD** | 0 (0%) | 0 (0%) | 0 (0%) | 0 (0%) | 0 (0%) | NA | 0 (0%) |
| **Had CHD event** | 331 (9.4%) | 248 (7.3%) | 802 (11%) | 217 (7.1%) | 821 (31%) | 1565 (49%) | 195 (3.6%) |
| **Follow-up time (years)** | 8.57 (2.64) | 10.49 (2.7) | 13.87 (2.79) | 2.71 (0.58) | 17.26 (5.48) | 5.20 (2.27) | 6.23 (1.24) |

\*fasting measures; \*\*number in pravastatin arm of the PROSPER trial; NA, not available.

**Figure 1:**
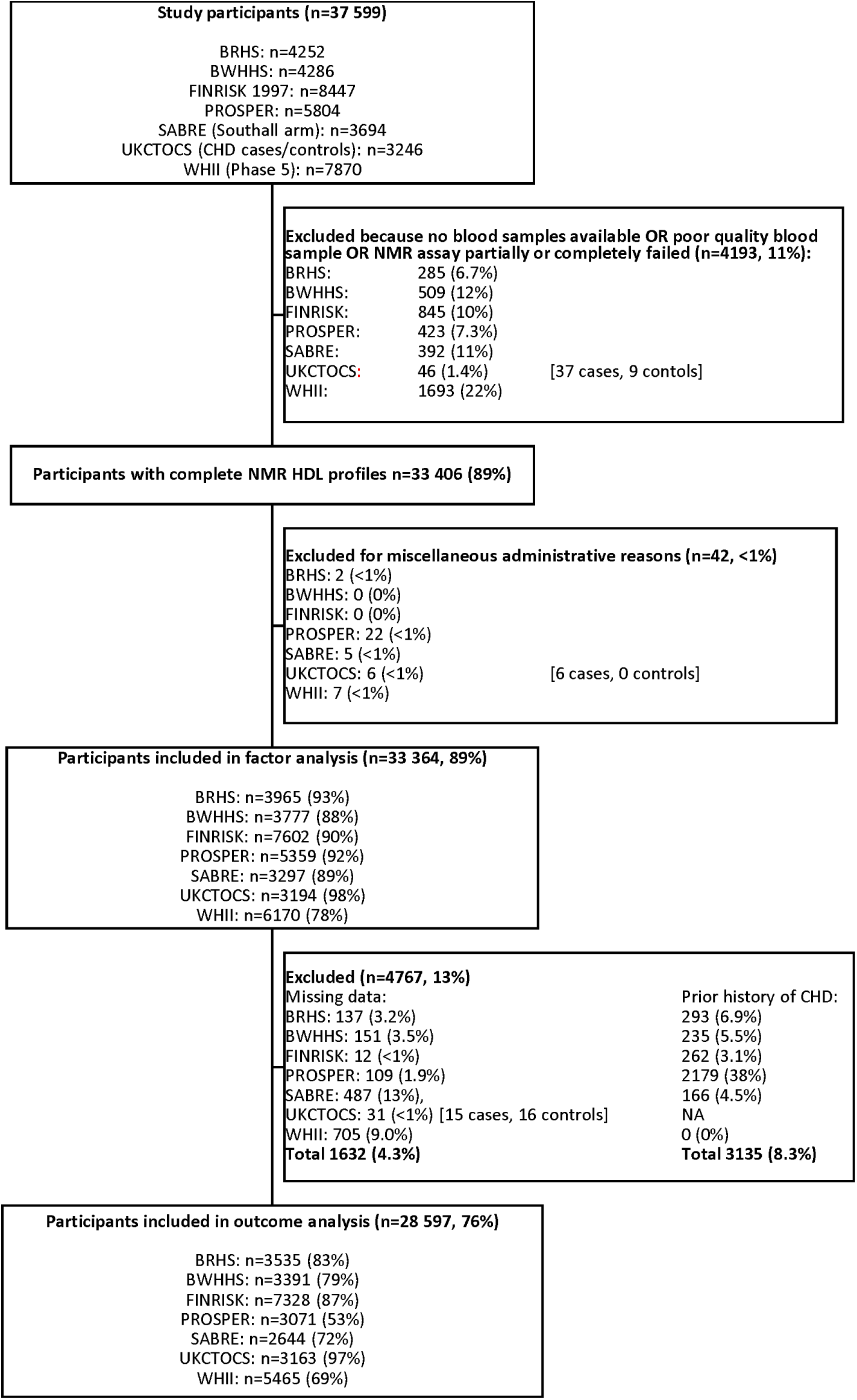
Participant flow for each study.

### HDL particles analysis using NMR spectroscopy

Serum blood samples were collected and stored after fasting in all studies according to study protocols, except those in FINRISK, which were after absorption was likely to be complete (median 5 hrs; 5.2% <4 hrs after last meal), and UKCTOCS, which were not taken in the fasting state. Biomarkers were quantified using high-throughput NMR spectroscopy (Nightingale Health Ltd, Helsinki, Finland), which provides simultaneous quantification of routine lipids, lipoprotein subclass profiling with lipid concentrations within 14 subclasses, fatty acid composition, and various low-molecular weight metabolites including amino acids, ketone bodies and glycolysis-related metabolites in molar concentration units. Details of the experimentation and epidemiological applications of the NMR platform are described elsewhere^16, 24^.

We selected 21 HDL biomarkers from the NMR output for this analysis, including lipoprotein components (free cholesterol (FC), cholesterol esters (CE), phospholipids (PL) and triglycerides (TG)) in each size class (extra-large (XL), large (L), medium (M) and small (S)), the concentration of particles circulating in each size class and the mean diameter of all HDL particles (**eTable 2**). We excluded HDL biomarkers that represent the direct sum of other measured lipids and derived biomarkers such as fractions.

### Ascertainment of coronary heart disease

Incident CHD events were defined as non-fatal myocardial infarction (MI) and fatal cardiac death in all studies. Some studies additionally included coronary artery by-pass (CABG), percutaneous transluminal coronary angioplasty (PTCA), unstable angina and definite angina. Outcome definitions used in each study are in **eTable 1**.

### Statistical analysis

We employed a two-stage analysis to investigate the association of specific components of the HDL pathway with incident CHD. This approach was applied independently in each study using individual patient data; see **Online-only material** for full technical details.

In the first stage we used unsupervised factor analysis to estimate ‘latent’ (i.e. unobserved but inferred through statistical modelling from observed variables) HDL factors as summary measures that attempt to capture systematic information from the correlation matrix of the 21 biomarkers. We fitted a saturated 15-factor model and, to determine the optimal number of factors, looked for eigenvalue >1 (**Figure 2**) and a threshold that provided a consistent solution across studies and was supported by plausible biological interpretations of the factors generated. We used maximum likelihood estimation followed by oblique quartimin rotation and defined resultant loadings ≥0.4 (in absolute value) as salient^25^. We assessed consistency of between-study solutions with a coefficient of variation (CV). From the optimal solution we predicted factor scores for individuals using the regression method^25^.

**Figure 2:**
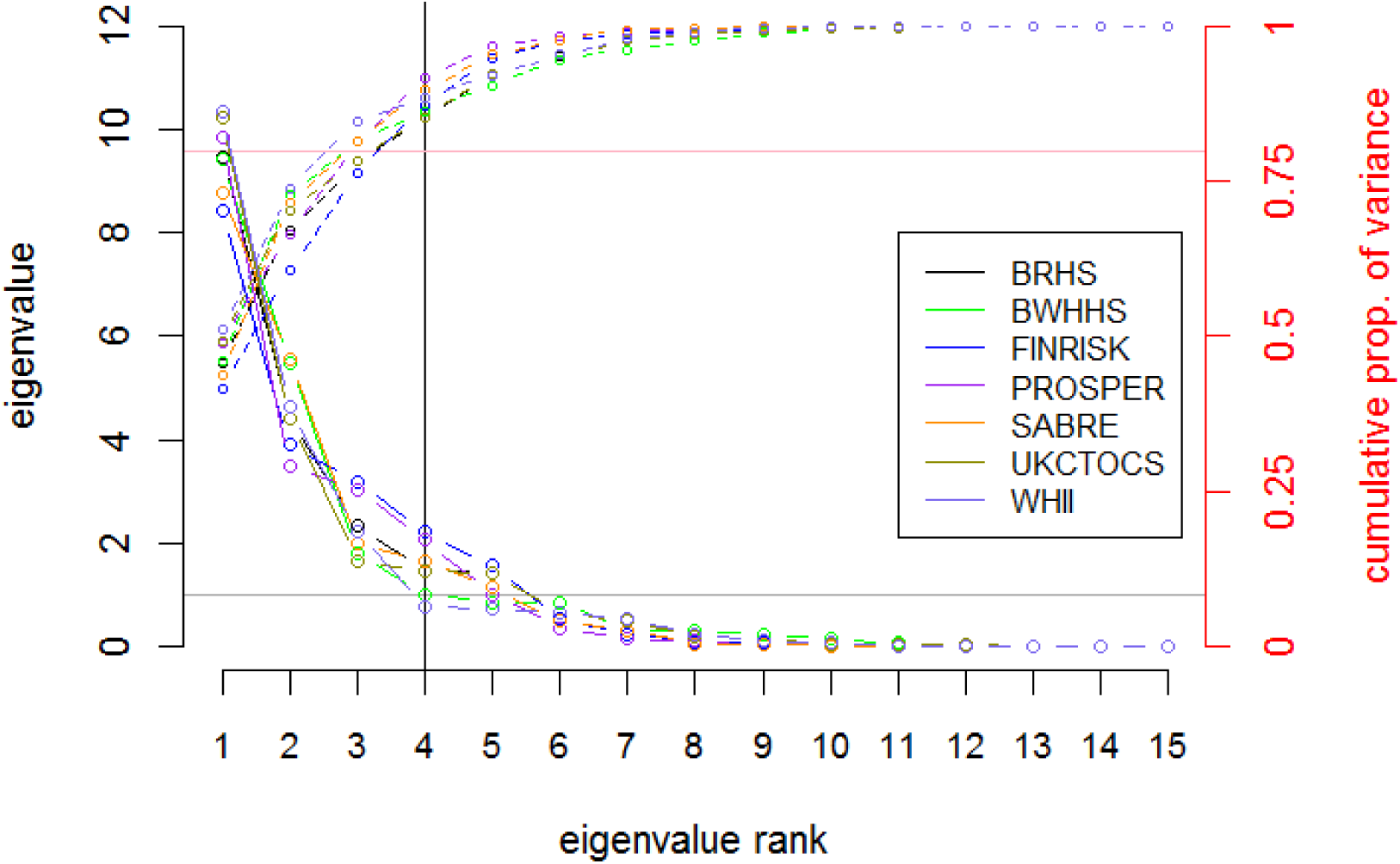
Screeplot of eigenvalues (solid lines, left y-axis) and cumulative proportion of variance explained (dashed lines, right y-axis) for each factor from the saturated 15-factor factor analysis solutions in seven studies. The grey and pink horizontal lines denote an eigenvalue of 1 and cumulative variance explained of 80% respectively. The vertical line indicates the optimal number of factors with most eigenvalues greater than 1 and >80% of variance explained.

In the second stage we explored the relationship between HDL factor scores and incident CHD in participants free of CHD at baseline in a complete-case analysis with Cox regression. Scores from each HDL factor were standardized (mean=0, SD=1). We evaluated factors jointly in a model adjusted for age, then progressively introduced confounders of CHD risk: sex, ethnicity, smoking, systolic blood pressure (SBP), body mass index (BMI), prevalent Type-2 diabetes mellitus, and LDL-C where available. We further adjusted for TG not carried by HDL as a sensitivity analysis; we use non-HDL-TG rather than total-TG to avoid collinearity with the HDL-TG present in both. We did not adjust for HDL-C, as its components (free-cholesterol and cholesterol-ester within each particle size) are already included in factor scores. In each study we excluded covariates with >10% missingness from modelling.

We pooled study-specific regression results in a random-effects meta-analysis using the DerSimonian-Laird estimator of between-study variability, and quantified heterogeneity with the I^2^ statistic^26^.

We used R (R Foundation for Statistical Computing, Vienna, Austria; http://www.r-project.org) for all statistical analyses; version 3.0.2 for meta-analysis.

## Results

A total of 33,364 subjects (47% women) with a mean age of 59.6 years contributed data to the factor analyses. In the regression analysis of the associations between factor scores and incident CHD, we excluded 1632 (4.3%) due to missing covariate or outcome data and 3135 (9.4%) due to history of CHD at or prior to baseline. Baseline characteristics of the participants and proportion of missingness for data used in both analyses are shown in **Table 1** and **eTables 3** and **4**. After a follow-up for CHD that ranged from a mean of 2.7 years in PROSPER to 17.3 years in SABRE, 4179 incident CHD events were accrued among the 28,597 participants with complete data and no history of CHD at baseline.

### NMR spectroscopy vs. enzymatic HDL-C measurements

We found the degree of correlation within studies between levels of HDL-C determined using enzymatic methods versus those determined using NMR spectroscopy to be between 0.76 and 0.90. There was good agreement between each one’s association with CHD in all studies with the two HRs varying by <15% (eTable 5).

### HDL factors

The pattern of correlation among HDL biomarkers was consistent across studies (**eFigure 1**) as was the distribution of HDL-C subfractions across total HDL-C (**eFigure 2**). Screeplots for all seven studies suggested the presence of 3 to 5 HDL factors (Figure 2). Content from the 4-factor solution was the most consistent between studies (**Figure 3** and **eFigure 3**). Each study estimated: a factor denoted extra-large (XL) that, when factor loadings were averaged, loaded highly on the 5 XL-HDL biomarkers (particle concentration, phospholipids, triglycerides, and free and esterified cholesterol) and the mean HDL particle diameter; a large (L) factor that loaded highly on the 5 L-HDL biomarkers and the mean HDL particle diameter; a medium/small (MS) factor that loaded highly on the 5 M-HDL biomarkers and 4 S-HDL biomarkers (excluding triglycerides); and a triglyceride (TG) factor that loaded highly only on triglycerides in all particle sizes.

**Figure 3:**
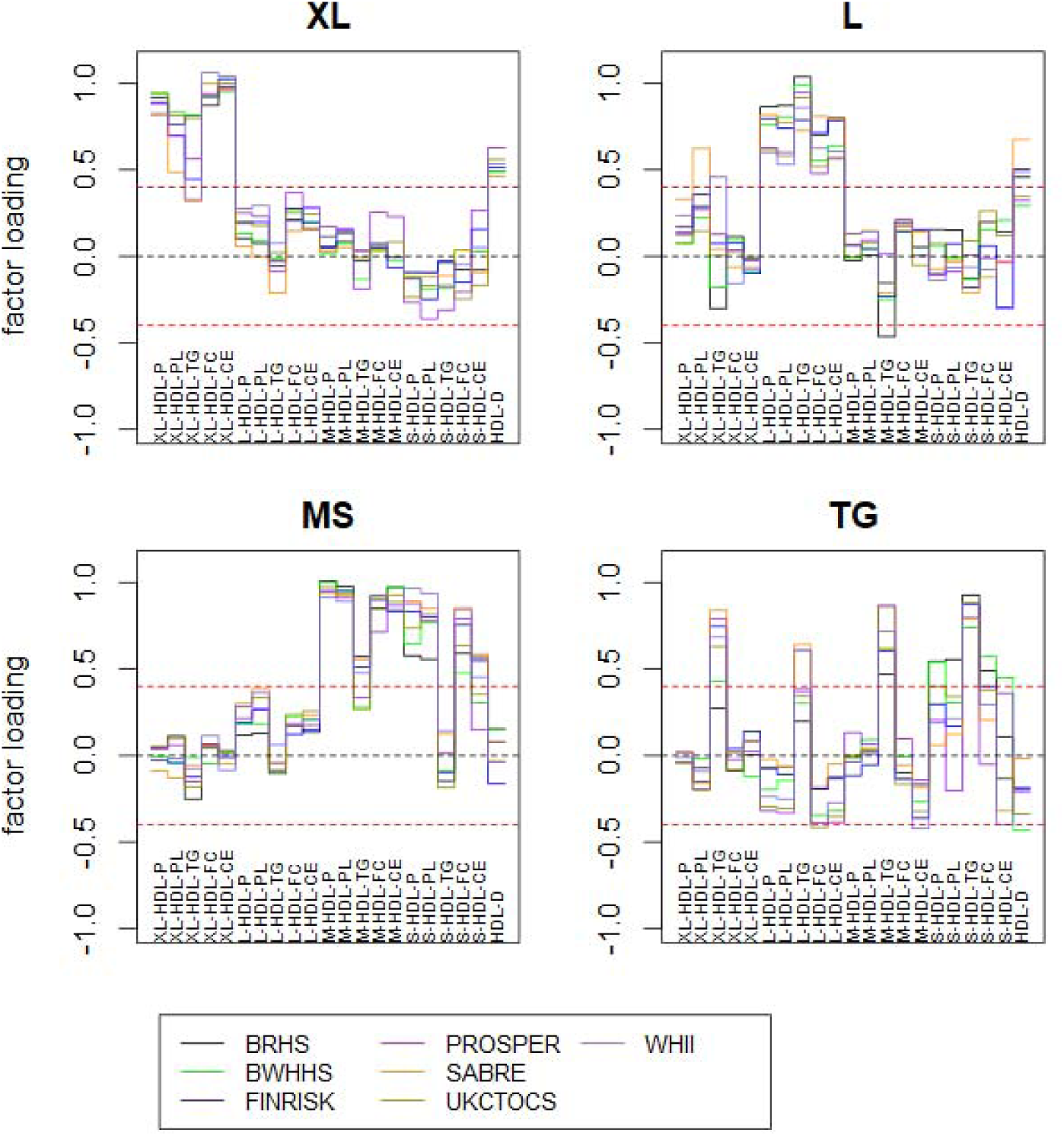
HDL biomarker factor loading patterns for 4 latent factors for each of the seven studies. Biomarkers with factor loadings outside the range of +/-0.4 factor loading (as indicated by the dotted lines) are considered ‘high’ loading.

The pattern of factor loadings was highly consistent between the seven studies: the median CV within each factor was 5.6, 10.1, 10.2 and 13.1% in XL, L, MS and TG respectively (**eTable 6**). The solution was also highly specific, with 17 out of 21 HDL biomarkers being associated with only one factor, even though in an unsupervised factor analysis they could load to all factors. Exceptions were the mean HDL diameter (associated with the XL and L factors) and triglycerides components in XL-, L- and M-HDL particles (associated with their relevant size factor and with the TG factor).

We observed moderate correlations between the XL and L factors (mean r=0.58) and between the L and MS factors (mean r=0.36). All other correlations among factors were <0.2; in particular, the MS factor did not correlate with XL (mean r=0.06) or TG (mean r=0.02). Correlation patterns were highly consistent across studies, with all CVs <12% (**eTable 7**).

### HDL factors vs. HDL-C

HDL-C was positively correlated with the XL (mean r= 0.67), L (r=0.79), and MS (r=0.63) HDL factors. These were driven by the correlations of HDL-C sub-fractions with their respective factors. HDL-C was negatively correlated with the TG factor (r=-0.32) (**eFigure 4**). Despite these moderate to strong correlations, HDL cholesterol sub-fractions make up less than half of the HDL factor scores: 42% of XL; 32% of L; 42% of MS and 25% of TG (**eFigure 5 and eTable 6**).

**Figure 4:**
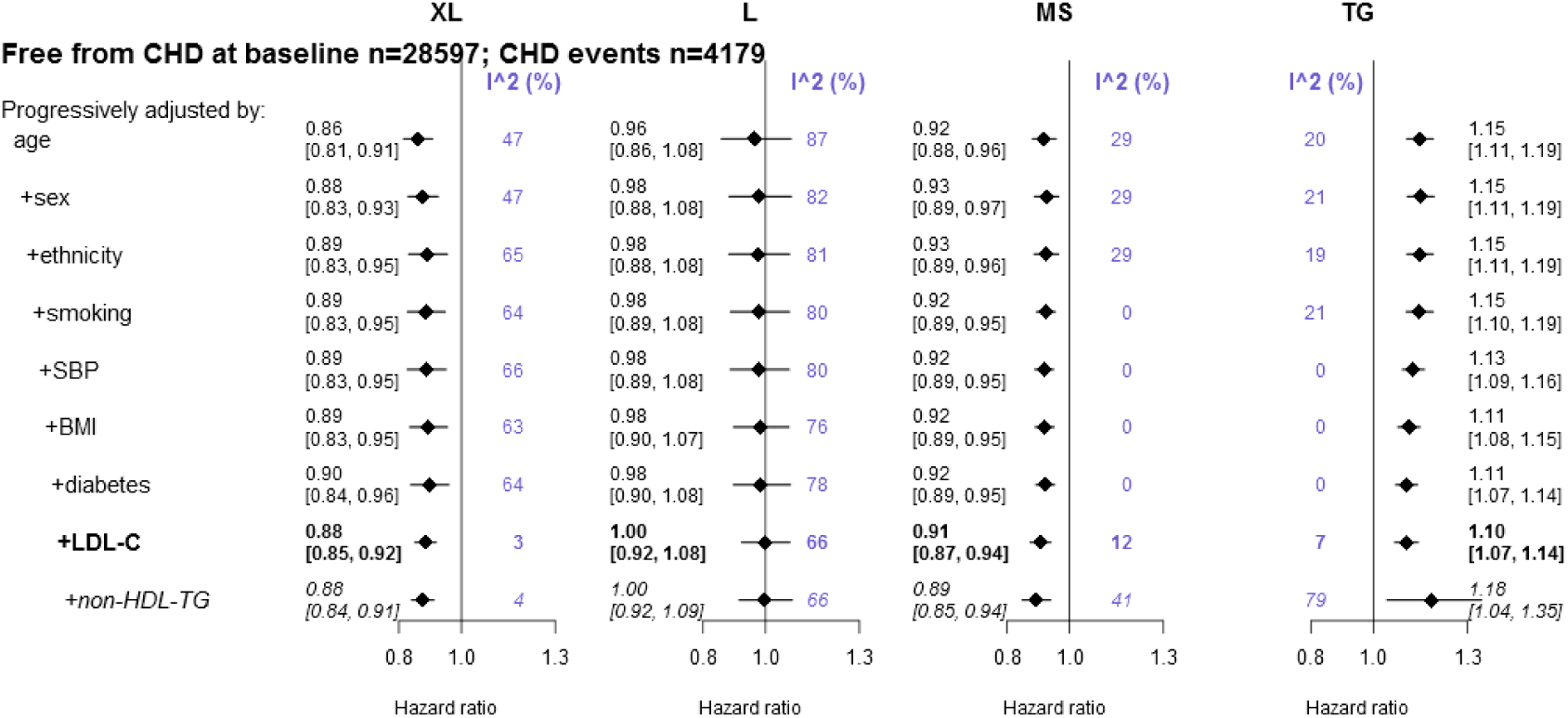
Associations between HDL factors and the risk of CHD in 28 597 participants with no history of CHD at the time of serum sampling. The factors are jointly modelled and minimally adjusted for age, then cumulatively by sex, ethnicity, smoking, systolic blood pressure, body mass index, diabetes, LDL-cholesterol concentration and non-HDL triglyceride concentration. Estimates represent the effect for a 1-SD increase in the normalised factor scores.

Where data was available we examined correlations between the HDL factors and HDL_2_-C, HDL_3_-C and apolipoprotein-B (**eTable 8**). HDL_2_-C was strongly correlated with the XL, L and MS factors (mean r=0.56, 0.76 and 0.71) and HDL_3_-C moderately so (r=0.45, 0.49 and 0.36). With the TG factor, they were correlated in opposite directions (HDL_2_-C: -0.39, HDL_3_-C: 0.24). These findings indicate a lack of specificity between HDL_2_-C and HDL_3_-C and the HDL factors. Apolipoprotein-B was uncorrelated with the XL and MS factors (r<0.01 and r=-0.02, respectively), but moderately negatively correlated with the L factor (r=-0.23) and positively correlated with the TG factor (r=0.57).

### Association between HDL factors and CHD

In 5 out 7 studies all potential confounders were available. Due to unavailability or missingness >10% UKCTOCS did not adjust for smoking, SBP and diabetes and WH-II did not adjust for BMI. Details of how we accounted for this in the meta-analysis are available in the **Online-only Material**. UKCTOCS did not record whether participants had a history of CHD prior to the time of serum sampling, so such participants (if any) could not be excluded or adjusted for and were included in the analysis.

**Figure 4** describes the summary results of the progressively adjusted Cox models on the four HDL factors. After adjusting for age and the other three factors an SD increase in the XL factor was associated with a 14% reduction in the risk of CHD (HR 0.86, 95% CI [0.81,0.91]), in the MS factor with an 8% reduction (0.92 [0.88,0.96]) and in the TG factor with a 15% increase (1.15 [1.11,1.19]). The progressive adjustment of established confounders for CHD (sex, ethnicity, smoking, SBP, BMI, diabetes, LDL-C) had minimal effect on these estimates and reduced between-study heterogeneity. After all adjustments there was evidence of 12% (0.88 [0.85, 0.92]) and 9% (0.91 [0.87, 0.94]) reduction in CHD risk per SD increase in the XL and MS factors and a 10% (1.10 [1.07, 1.14]) increase in risk per SD increase in the TG factor, with little heterogeneity between studies (all I^2^≤12%). There was no evidence of an association between the L factor and CHD (1.00 [0.92, 1.08]; I^2^=66%). **eFigure 6** shows study-specific HRs for the minimally- and maximally-adjusted models.

Associations of the XL, L and MS factors with CHD did not change after further adjustment by non-HDL triglycerides. However, the TG factor lost a substantial amount of precision, likely due to collinearity between this factor and TG in other lipoproteins, which were strongly correlated in most studies (range: 0.39, 0.88). Our results were unaltered by excluding data from the statin arm of PROSPER.

## Discussion

In this analysis including up to 33,364 individuals and 4,197 coronary events, we provide evidence indicating heterogeneous associations between components of the HDL-pathway, proxied through four HDL factors, with the risk of CHD. The HDL components represented by HDL biomarkers covering size and composition on extra-large and medium/small particles were inversely associated with CHD, while the triglyceride component was directly associated after adjustment for potential confounders including LDL-C. No association was observed for the component represented by biomarkers covering size and composition of large particles.

The potential benefits of the HDL-components we identified, compared with HDL-C, can be illustrated through an analogy on total-cholesterol versus LDL-C. Initial technology suitable for population-based studies was capable of capturing only total-cholesterol, and this measurement, in studies like the Framingham Heart Study^27^, showed a harmful association with CHD. Improvements in technology made the measurement of major cholesterol sub-fractions possible and revealed opposite associations: LDL-C was harmful but HDL-C was protective. In this study, further improvements in technology through NMR spectroscopy allowed us to uncover heterogeneous associations within the HDL pathway.

The four HDL components described in this analysis were highly reproducible across seven studies, despite important differences in study design and participant characteristics. This high reproducibility suggests these factors are very likely to have distinctive biological determinants. We anticipate that discovery of their biological determinants using genomics, proteomics and functional assays could help refine selection of intervention targets. Given the qualitatively different associations with CHD, genomic studies aimed for target selection should consider the potential associations that each gene-encoded protein may have on four HDL components modelled jointly.

Until now, target-selection, compound development, early pre-clinical and randomized trials have used HDL-C as the sole metric to drive the drug-discovery process. With this approach, if two distinct drug targets (and associated compounds) had a similar effect on HDL-C concentration, they would be expected to have similar therapeutic efficacy against CHD. However, evidence from large-scale phase-3 trials, other observational studies showing that HDL-C protection disappears with adjustment for other HDL particles^28, 29^, and the results we present in this paper do not support this hypothesis.

Additional support for the HDL factors we identified comes from the good agreement with the five HDL subclasses (extra-large, large, medium, small and very small) proposed by Rosenson et al^13^. These categories were proposed as a new nomenclature to capture the diverse data around HDL physical properties derived from various HDL measurement techniques, but not including the specific NMR platform used in this report. Although previous studies^28-30^ have attempted to uncover the impact of HDL sub-classes on CHD using NMR, our study provide novel lines of evidence. First, previous studies only measured HDL particle size and not composition within HDL sub-classes. Therefore, their results do not truly capture the impact of HDL as a pathway. An example of this is that role of triglycerides carried only within HDL particles was not assessed in previous studies. Second, our study covering seven cohorts includes ten times more CHD cases than any of previous studies, refuting any concern on inconsistency of findings. Taking into consideration those differences, our findings also mirror those from the VA-HIT trial, which found that medium and small HDL particles were protective against CHD while the large particles were not. Our analysis has several strengths. First, we observed reproducible estimates for the HDL components in independent, unsupervised factor analyses across seven diverse study populations. Second, despite using an unsupervised strategy that allows each biomarker to associate with all factors, the final composition of the factors had high specificity (17 of 21 loading on a single factor) and strong biological plausibility. For example, the biomarkers that formed the XL, L and MS factors were largely represented by particle concentrations, phospholipids, free-cholesterol and cholesterol esters from their respective HDL-subclasses. Third, we observed minimal between-study heterogeneity in regression estimates for three of the four factors. Fourth, several publications^31-33^ have shown the validity of the NMR platform used in this analysis. For example, genetic determinants of major lipid fractions measured through NMR are consistent with those discovered by enzymatic methods^31^. We also showed strong correlation between NMR HDL-C and enzymatic HDL-C and concordant CHD protective associations across all studies.

We chose not to use total HDL-C for the construction of the factors. Instead we used their components and hence did not adjust estimates for total HDL-C, as the variables that comprise it were already included in the factors. Adjustment would be desirable if the aim was to propose to use these new HDL factors as better measures of “risk prediction” for CHD. Instead our goal was to identify components of the HDL pathway relevant for drug-discovery. By analogy, total cholesterol, instead of LDL-C, is the biomarker used in “risk prediction” models like Framingham^34^ or the Pooled Cohort Equations^35^, despite being of less value for drug-discovery.

Several findings indicate that the HDL factors are distinct from cholesterol carried across HDL particles of different size. First, free-cholesterol and cholesterol ester within HDL particles accounted for less than half of the value of HDL factor scores. Second, HDL_2_-C and HDL_3_-C were not strongly correlated with the HDL-factors. Third, despite HDL-C having moderate-to-strong correlations with the HDL-factors, the factors that showed associations with CHD were largely uncorrelated between themselves.

Even though we had access to a large sample and standardized analysis plan, our results need to be replicated in other large-scale surveys such as the UK-Biobank or the Mexico City Prospective study where the same NMR platform-based output should be available in the near future. This increased sample size will allow robust detection of small associations such as those we report in the current study and avoid use of heterogeneous datasets in terms of study design, outcome ascertainment and missingness and avoid the need for assumptions related to meta-analytic techniques. Additional advantages of studies like UK-Biobank^36^ and INTERVAL^37^, with genomics and serum NMR measurements, are the possibility to explore if the associations of HDL factors with incident CHD are likely to be causal.

In conclusion, our HDL factor analysis uncovered the presence of heterogeneous associations between HDL-pathway factors and CHD and suggests that the full biological effect of these pathways is unlikely to be captured by HDL-C alone. If our findings are validated in independent cohorts in future analyses using genetics and proven to be causal, this will help to precisely inform the selection of drug targets and to guide the drug discovery process from pre-clinical phase to phase-3 trials.

## Data Availability

Data access is conditional on individual study approval in accordance with existing ethics and governance rules for each study. Summary level data may be made available on request subject to the approval of individual studies.

## Acknowledgements

The authors acknowledge and thank the participants and study teams from BRHS, BWHHS, FINRISK, PROSPER, SABRE, UKCTOCS and WH-II for data collection and support and Drs Reecha Sofat and Rui Providencia for providing valuable comments on the manuscript.

